# Improved specificity of glutamate decarboxylase 65 autoantibody measurement using luciferase-based immunoprecipitation system (LIPS) assays

**DOI:** 10.1101/2023.07.03.23292157

**Authors:** R.C. Wyatt, S.L. Grace, C. Brigatti, I. Marzinotto, B.T. Gillard, D. Shoemark, K. Chandler, P. Achenbach, L. Piemonti, The BOX Study Group, A.E. Long, K.M. Gillespie, V. Lampasona, A.J.K. Williams

## Abstract

Autoantibodies to glutamate decarboxylase (GADA) are widely used in the prediction and classification of type 1 diabetes. GADA radiobinding assays (RBAs) using N-terminally truncated antigens offer improved specificity but radioisotopes limit the high-throughput potential for population screening. Luciferase-based Immunoprecipitation System (LIPS) assays are sensitive and specific alternatives to RBAs with the potential to improve risk stratification.

The performance of assays using the Luciferase (Nluc-) conjugated GAD_65_ constructs, Nluc-GAD65(96-585) and full length Nluc-GAD65(1-585) were evaluated in 434 well-characterised sera from recent-onset type 1 diabetes patients and first-degree relatives.

Non-radioactive, high-throughput LIPS assays are quicker and require less serum than RBAs. Of 171 relatives previously tested single autoantibody positive for autoantibodies to full-length GAD_65_ by RBA but had not progressed to diabetes, fewer retested positive by LIPS using either truncated (n=72) or full-length (n=111) antigen. The Nluc-GAD65(96-585) truncation demonstrated the highest specificity in LIPS assays overall but in contrast to RBA, N-terminus truncations did not result in a significant increase in disease-specificity compared with the full-length antigen. This suggests that binding of non-specific antibodies is affected by the conformational changes resulting from addition of the Nluc antigen. Nluc-GAD65(96-585) LIPS assays offer low blood volume, high specificity GADA tests for screening and diagnostics.

## Introduction

Approximately 80% of patients with type 1 diabetes are positive for autoantibodies to glutamate decarboxylase (GADA)(1). This test is widely used in prediction and classification of diabetes.

Commonly used methods for measuring GADA include RBA(2) and ELISA(3). Despite efforts to improve GADA measurement, many testing single GADA positive are unlikely to develop diabetes(4). There is consensus that diabetes-associated GADA primarily recognise epitopes in the middle and C-terminal regions of GAD_65_, while autoantibodies specific to the N-terminal have little association with progression(5). We previously showed that the first 142 amino acids of GAD_65_ do not contribute to epitopes recognised by disease-associated autoantibodies(6). Using N-terminally truncated GAD_65_ radiolabels ^35^S-GAD_65_(98–585) and ^35^S-GAD_65_(143–585) improved the specificity of RBAs without impacting sensitivity. In first-degree relatives(FDR) of patients, autoantibodies measured with these constructs were more closely associated with diabetes risk(7).

However, the use of radioactive tracers in RBAs is costly and time-consuming with environmental implications; moreover, their future availability is in question which potentially limits the long-term sustainability of RBA. Alternative methods including electrochemiluminescence(ECL), antibody detection by agglutination-PCR(ADAP), bridge ELISAs, chemiluminescence immunoassays(CLIA) and Luciferase-based Immunoprecipitation System(LIPS) assays are increasing in popularity(8). Good performance of LIPS assays to measure type 1 diabetes associated insulin(IAA)(9) and islet-antigen 2(IA-2A)(10) autoantibodies has been reported.

Using well-characterised samples from the Bart’s Oxford (BOX) family study(11), we evaluated how GADA levels measured by LIPS, using full-length and truncated antigens, compared with the equivalent RBAs.

## Research Design and Methods

### Population

#### Screening cohort

To assess sensitivity and specificity of the Nluc-GAD_65_ constructs we selected 11 patients with recent-onset type 1 diabetes [5 (45.5%) male; median duration 43 days (range-1 to 84 days)] and 25 low-risk GADA(96-585) positive first-degree relatives (FDRs) of patients [8 male (32%)]. FDRs were considered low risk as they had not developed diabetes during follow-up [median 19 years (range1.2-29.7 years)] and did not have additional islet autoantibodies. These samples had been tested previously by RBA using full-length GAD_65_ and five truncated GAD_65_ constructs(6).

#### Evaluation cohort

Samples were selected for detailed evaluation of the LIPS assay, based on previous studies(6, 7) including 156 patients with recent-onset type 1 diabetes (2 had insufficient sera and were excluded) and 740 FDRs followed for disease development by questionnaire (Table 1).

Table 1

**Table 1**
|  | Patients<br>(n=154) | GADA Positive Relatives<br>(n=271) |  | GADA Negative Relatives<br>(n=461) |  |
| --- | --- | --- | --- | --- | --- |
|  |  | Progressor<br>(n=64) | Non<br>Progressor<br>(n=207) | Progressor<br>(n=32) | Non<br>Progressor<br>(n=429) |
| Males | 94<br>(60%) | 31<br>(48%) | 100<br>(48%) | 23<br>(72%) | 205<br>(48%) |
| Median age, years (range) | 11.7<br>(1.3-20.9) | 33.3<br>(5.7-52.9) | 30.6<br>(1.33-57.4) | 39.2<br>(1.4-56.2) | 32.6<br>(1.7-57.3) |
| Median age at diagnosis,<br>years (range) | 15.3<br>(2.7-100) | 37.5<br>(11.7-69.8) | - | 48.3<br>(5.1-68.5) | - |
| Median follow-up,<br>years (range)/<br>Median diabetes duration,<br>days (range) | 1<br>(-7-90) | 7.56<br>(0.2-27.8) | 17.6<br>(0.6-30.8) | 12.8<br>(1.6-23.8) | 17.6<br>(0-31.0) |
| Additional autoantibodies* | 140 | 34 | 37 | 4 | 11*** |
| IA-2A | 120 | 19 | 13 | 1 | 1 |
| IAA | 71** | 19 | 23 | 3 | 10 |
| ZnT8A | 111 | 19 | 17 | 0 | - |

Data are number of first-degree relatives (%) or median (range).
\*Additional autoantibodies are from IA-2A, IAA and ZnT8A.
\*\*57 patients not tested for IAA as the sample was taken more than two weeks after diagnosis and any antibody to insulin may derive from insulin therapy induction.
\*\*\*Not tested for ZnT8A

Of 740 FDRs, 278(37.6%) were previously found GADA positive by radioimmunoassay. Of these, 69(25%) developed diabetes, 254(91%) had follow-up data for survival analysis and 7 with insufficient sera and were excluded. The remaining 462 FDRs previously tested GADA negative. This population was enriched with 32(6.9%) FDRs who developed diabetes (excluding one with insufficient sera (Table 1)).

All samples were previously tested for GADA using the harmonised RBA protocol with the full-length ^35^S-GAD_65_(1-585) and N-terminally truncated ^35^S-GAD_65_(96-585) antigens(7). Data on additional autoantibodies [IA-2A, IAA and zinc transporter 8 autoantibodies (ZnT8A)] were also available.

#### Recombinant luciferase tagged GAD_65_ antigen production

Comparable to the harmonised RBA protocol(12) recombinant Nanoluc® luciferase (Nluc) tagged GAD_65_ antigens (Figure 1a) were encoded in a pCMVTnT™ plasmid (Promega, USA) and synthesised using the SP6 TnT quick coupled *in vitro* transcription/translation kit (Promega) using 1microgram of antigen and a 1.5hr incubation at 30°C. The antigen was purified using a NAP5™ desalting column (packed with Sephadex™ G-25 – Illustra, supplied by VWR, UK) and Tris-buffered saline with Tween-20 buffer (TBST; 20 mM Tris, 150 mM NaCl, pH 7.4, and 0.5% Tween-20), by collecting 3 fractions (400, 200 and 500µl). Luciferase activity was quantified in light-unit equivalents (LU) by measuring the emitted bioluminescence of 2µl of antigen mixed with 40µl of Nano-Glo® substrate (as per manufacturer instructions, Promega) in a Centro XS3 luminometer (Berthold Technologies GmbH&Co. Germany) for 2 seconds/well. A typical reaction yielded between 10^6^-10^7^ LU/µl of antigen in a total volume of 600µl pooled from the first two fractions. The antigen was divided into aliquots and stored at –70°C.

**Figure 1.**
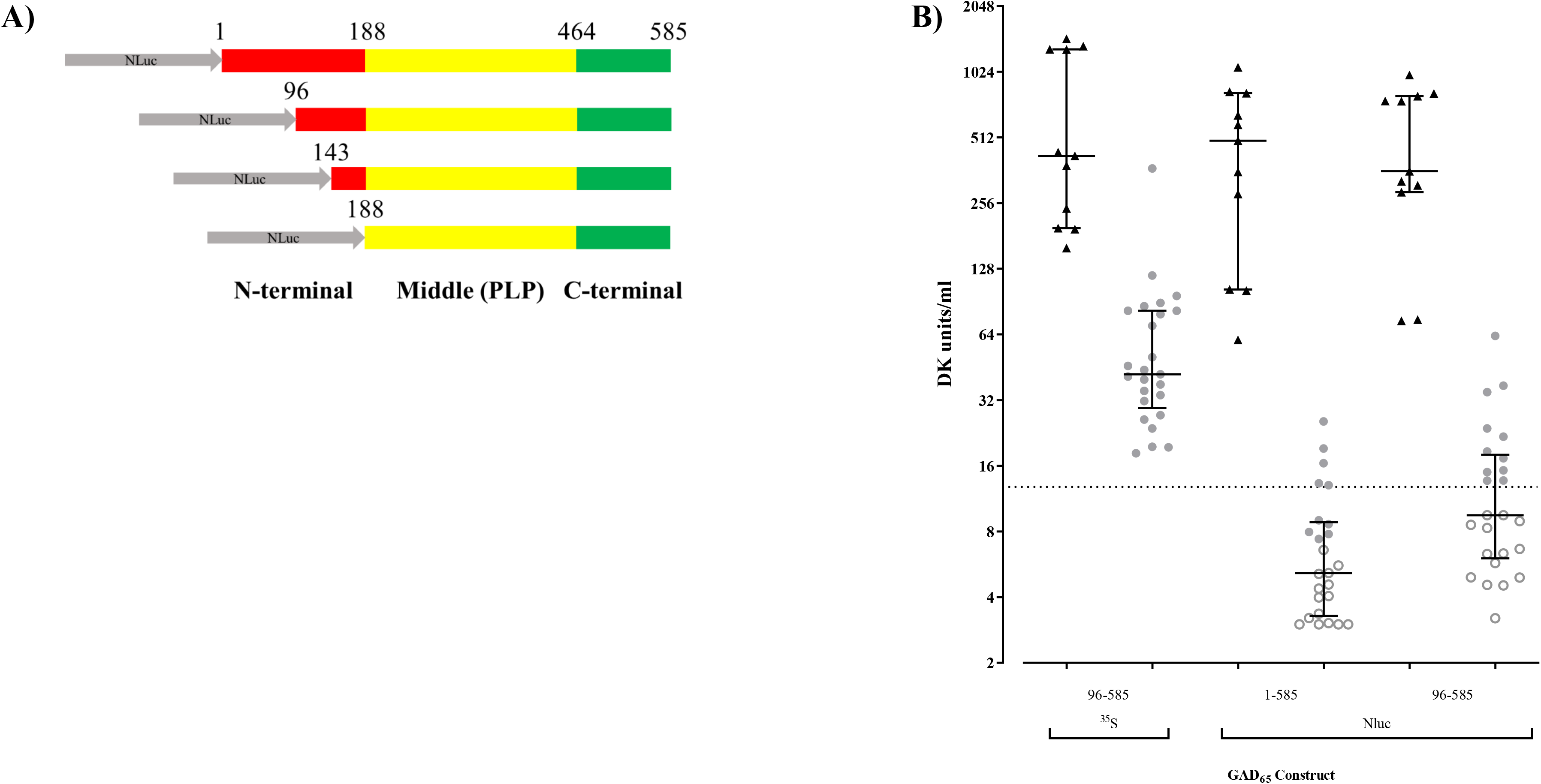
(a) Diagram of the NanoGlo® luciferase (Nluc) GAD_65_ constructs which were assessed for the sensitivity and specificity of GADA measurement by Luminescence Immunoprecipitation System (LIPS) assay. (b) A plot of 25 low-risk single GADA(96-585) positive relatives who had not developed diabetes during follow-up (grey circle) and 11 recent-onset T1D patients (black triangle) who were measured for GADA using ^35^S-GAD_65_(96-585) in radioimmunoassay and Nluc-GAD_65_(1-585) & Nluc-GAD_65_(96-585) in LIPS assays. Filled triangles and circles indicate positives by that construct by individual assay thresholds. Dotted line indicates positivity threshold for ^35^S-GADA(96-585). In patients, the median antibody levels (DK units/ml) for ^35^S-GADA(96-585), Nluc-GADA(1-585) and Nluc-GADA(96-585), were 341.29 (range, 157.2-1267.3), 494.4 (range, 60.4-1072.5) and 357.8 (range, 73.8-988.1), respectively. In relatives, the median antibody levels were 42.0 (range, 18.3-368.5), 5.16 (range, 0.39-25.6) and 9.5 (range, 3.2-63), respectively.

#### GADA LIPS assay

The Nluc-GAD_65_ antigen was diluted in TBST + 0.1%BSA, to a concentration of 4.0×10^6^ LU/25µl (±200,000 LU). Sera (1µl, 2 replicates) were pipetted into a 96-well plate (Sarstedt, Nümbrecht, Germany) and incubated with 25µl diluted Nluc-GAD_65_ antigen for 2.5hr at room temperature protected from light. Immunocomplexes were precipitated using a 25% Protein A Sepharose 4 fast flow (PAS) suspension [6.25µl/well washed four times in TBST + 0.1% BSA; GE Healthcare Life Sciences, Amersham, UK) and a 1hr incubation with orbital shaking (∼700 rpm) at 4°C. Unbound Nluc-GAD65(96-585) was removed by five serial washes with TBST each including centrifugation (500xg/4°C/3min) and automatic buffer removal and dispensing (BioTek Elx405, Agilent, USA). Resin pellets were transferred into 96-well OptiPlates™ (Perkin-Elmer, USA), centrifuged (500xg/4°C/3min) and aspirated to a final volume of 30µl. Nano-Glo® substrate (40µl) was injected into each well immediately prior to LU determination using a standardised protocol on the Centro XS3 luminometer (inject, shake 5 seconds/well, detect 2 seconds/well).

In the 2020 Islet Autoantibody Standardization Program (IASP) workshop, the adjusted sensitivity at 95% specificity (AS95) for GADA(1-585) and GADA(96-585) measured by radioimmunoassay was 78% and 84%, respectively. For GADA(1-585) and GADA(96-585) measured by LIPS, the AS95 was 76% and 86%, respectively.

#### Antibody quantification and thresholds

Logarithmic standard curves were generated from standards established from the National Institute of Diabetes and Digestive and Kidney (NIDDK) disease harmonisation programme, allowing quantification of autoantibody levels (DK units/ml)(12) Thresholds for GADA by RBA and LIPS were set at the 97.5^th^ percentile of 221 healthy schoolchildren. This was equivalent to 13.5, 12.8, 7.3 and 10.7 DK units/ml for ^35^S-GAD_65_(1-585), ^35^S-GAD_65_(96-585), Nluc-GAD_65_(1-585) and Nluc-GAD_65_(96-585), respectively.

#### Statistical Analysis

Wilcoxon matched-pairs signed-rank tests were used to compare antibody levels and McNemar’s test with Yate’s correction to compare antibody status with different GAD_65_ constructs and assay formats. Kaplan-Meier curves with Mantel-Cox log-rank test were used to compare survival between groups. For all analyses, a two-tailed P <0.05 was considered significant. The partial area (90th percentile) under the curve (pAUC) of the receiver operating characteristic (ROC) with 95% CI was calculated assuming a nonparametric distribution of results using R softwareV3.2.2. Other statistical analyses were performed using GraphPad PrismV6.

## Results

### Construct screening

GADA measured in sera from the screening cohort using LIPS assays with the four Nluc-GAD_65_ constructs (Figure 1a) were compared with those for GADA(96-585) obtained by RBA. Median antibody levels with the Nluc constructs were similar to ^35^S-GAD_65_(96-585) in patients (p>0.05 for all comparisons)(Figure 1b).

In 25 low-risk single GADA(by either RBA) positive relatives, median GADA levels, were lower when measured by LIPS than when measured by RBA with ^35^S-GADA(96-585) (p<0.01 for all comparisons)(Figure 1b).

Additional constructs tested (Nluc-GAD_65_(143–585) & Nluc-GAD_65_(188–585) did not improve discrimination further (data not shown). Therefore, Nluc-GAD_65_(1-585) and Nluc-GAD_65_(96-585) were selected for detailed evaluation compared with the highly diabetes-specific RBA using ^35^S-GAD_65_(96-585).

### Assay evaluation

#### The sensitivity of Nluc-GAD_65_ constructs were comparable to the higher specificity ^35^S-GAD_65_(96–585) construct in patients and high-risk relatives

Of 154 patients, 125(81%) were positive for ^35^S-GADA(1-585), and 125(81%) for ^35^S-GADA(96-585). More of these patients were positive for Nluc-GADA(1-585) than Nluc-GADA(96-585), 129(84%) vs. 116(75%), (p=0.0036). There was very good correlation between ^35^S-GADA(96-585) and Nluc-GADA(1-585) [r=0.96 (95% CI 0.94-0.97), p<0.0001] and Nluc-GADA(96-585) [r=0.93 (95% CI 0.90-0.95), p<0.0001] (Figure 2a).

**Figure 2.**
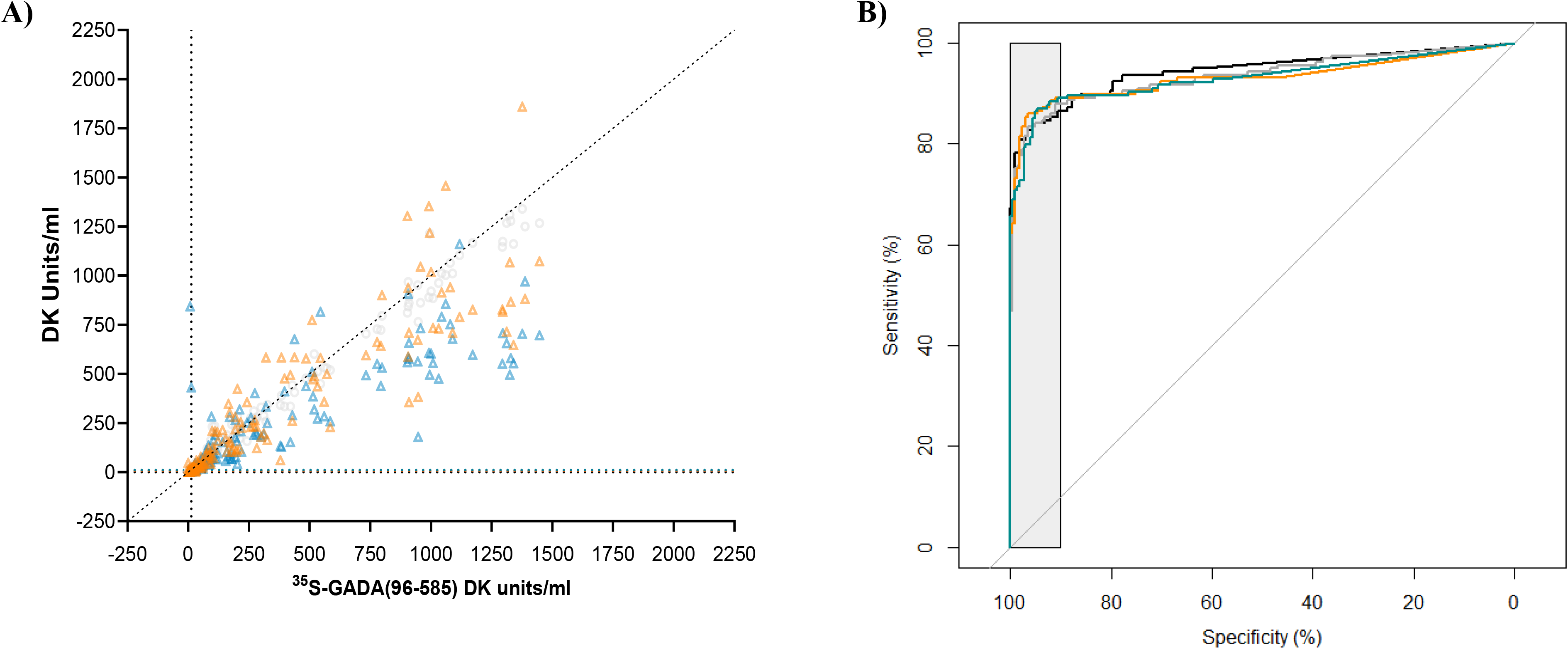
(a) A plot of ^35^S-GADA(1-585) (grey triangles), Nluc-GADA(1-585) (orange triangles) and Nluc-GADA(96-585) (teal triangles) levels against ^35^S-GADA(96-585) levels in 156 patients with recent-onset T1D. Overall, correlation of ^35^S-GADA(1-585), Nluc-GADA(1-585) and Nluc-GADA(96-585) with ^35^S-GADA(96-585) was excellent (r=0.99, 0.91 and 0.87, respectively. p<0.0001 for all). (b) Receiver operator characteristic curve for ^35^S-GADA(1-585) (black line), ^35^S-GADA(96-585) (grey line), Nluc-GADA(1-585) (orange line), and Nluc-GADA(96-585) (teal line) measured by radioimmunoassay or LIPS based on data from 156 patients with newly diagnosed T1D and 221 healthy schoolchildren. The area under the curve was 0.94 for ^35^S-GADA(1-585), 0.93 for ^35^S-GADA(96-585), 0.93 for Nluc-GADA(1-585) and 0.93 for Nluc-GADA(96-585). The partial AUC (at specificities >90%, within the grey box) was 0.082 for ^35^S-GADA(1-585), 0.081 for ^35^S-GADA(96-585), 0.084 for Nluc-GADA(1-585) and 0.082 for Nluc-GADA(96-585).

ROC analysis showed that Nluc-GADA(1-585) and Nluc-GADA(96-585) could discriminate between patients and healthy schoolchildren with comparable sensitivity and specificity to ^35^S-GADA(1-585) and ^35^S-GADA(96-585). Partial area under the curve values (at specificities >90%) were 0.082 (95% CI 0.075-0.088), 0.081 (95% CI 0.074-0.087), 0.084 (95% CI 0.077-0.089) and 0.082 (95% CI 0.076-0.088) for ^35^S-GADA(1-585), ^35^S-GADA(96-585), Nluc-GADA(1-585) and Nluc-GADA(96-585), respectively (Figure 2b).

Of 100 relatives who progressed to diabetes during follow-up and/or had autoantibodies to additional islet antigens, 84% were positive for all four specificities. Ninety-three (93%) were positive for ^35^S-GADA(1-585), 90% for ^35^S-GADA(96-585), 94% for Nluc-GADA(1-585) and 88% for Nluc-GADA(96-585) (ESM 1a).

#### Fewer single GADA positive relatives who have not developed diabetes were positive for GADA measured using LIPS assays

74(43%) of 171 GADA positive relatives who had no additional autoantibodies and had not progressed to diabetes during follow-up had antibodies to Nluc-GAD_65_(96-585). This was a lower proportion than for ^35^S-GADA(1-585) (156-90%; p<0.0001), ^35^S-GADA(96-585) (108-62%; p<0.0001) or Nluc-GADA(1-585) (112-65%, p<0.0001), respectively (ESM 1b).

Of the 430 relatives who originally tested GADA(1-585) negative and remained diabetes-free during follow-up 12(3%) were found positive for ^35^S-GADA(1-585), 6(1%, p>0.05) for ^35^S-GADA(96-585), 17(4%, p>0.05) for Nluc-GADA(1-585) and 10(2%, p>0.05) for Nluc-GADA(96-585) (ESM 1b).

#### Nluc-GAD65 antigens offer improved discrimination of risk in ^35^S-GADA(1-585) positive <u>relatives</u>

In 254 relatives, with follow-up data who tested GADA positive using the harmonised RBA with ^35^S-GAD_65_(1-585), the 15-year risk of diabetes was 25% (95% CI 20%-31%). Within this group, positivity for ^35^S-GADA(96-585), Nluc-GADA(1-585) and Nluc-GADA(96-585) further stratified risk of diabetes (p<0.0001, for all comparisons). Individuals positive for ^35^S-GADA(96-585) had a 32% (95% CI 25%-40%) risk of diabetes within 15 years, while individuals positive for Nluc-GADA(1-585) had a 30% (95% CI 24%-38%) risk and positivity for Nluc-GADA(96-585) had a 30% (95% CI 23%-39%) risk (Figure 3).

**Figure 3.**
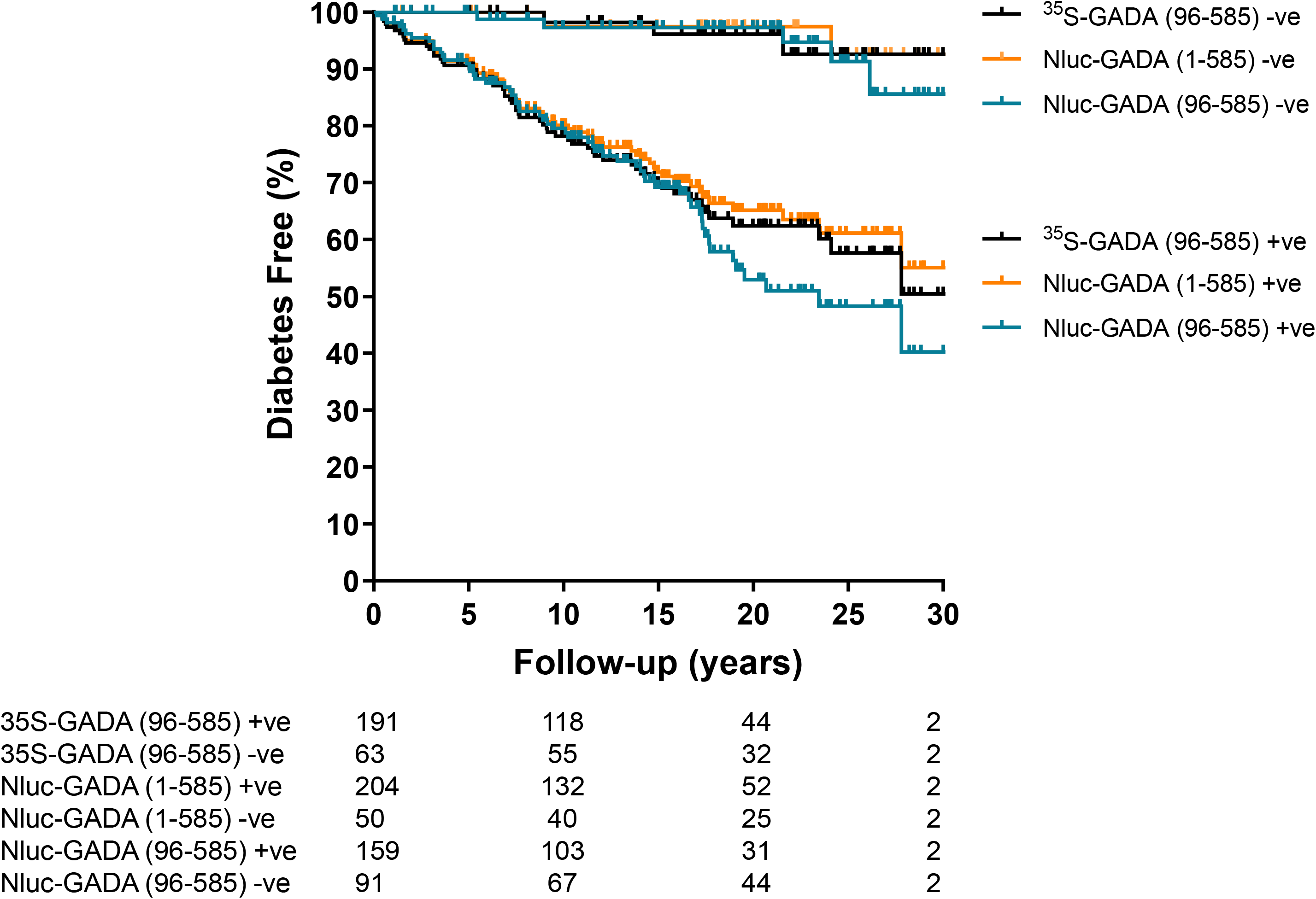
A Kaplan-Meier survival curve for first-degree relatives positive for ^35^S-GADA(1-585) according to positivity for ^35^S-GADA(96-585) (black lines), Nluc-GADA(1-585) (orange lines) and Nluc-GADA(96-585) (teal lines). ^35^S-GADA(96-585), Nluc-GADA(1-585) and Nluc-GADA(96-585) identified relatives at increased risk of diabetes progression. Individuals positive for ^35^S-GADA(96-585) had a 32% risk of developing diabetes within 15 years, while individuals positive for Nluc-GADA(1-585) or Nluc-GADA(96-585) had a 30% risk.

## Discussion

LIPS assays provide a high-performance alternative to well-established RBAs, widely used in diabetes studies. We compared LIPS Nluc full-length GAD_65_(1-585) and truncated GAD_65_(96-585) assays with the equivalent RBAs, particularly the truncated assay which demonstrated improved specificity in T1D and LADA(13, 14). Unexpectedly the full-length LIPS assay preformed almost as well as the truncated assays and much better than the equivalent RBA.

The sensitivity of GADA measurement by LIPS using Nluc-GAD_65_(1-585) and Nluc-GAD_65_(96-585) was comparable to RBAs using ^35^S-GAD_65_(1-585) and ^35^S-GAD_65_(96-585) in patients and high-risk relatives. The specificity of the LIPS assay using Nluc constructs was also improved compared with the harmonised RBA using ^35^S-GAD_65_(1-585) and/or ^35^S-GAD_65_(96-585). Relatives positive for GADA using LIPS were at increased risk of developing diabetes within 15 years versus those who were positive for GADA(1-585) by RBA but had a similar risk to those positive using ^35^S-GAD_65_(96-585). The sensitivity of GADA measurement was maintained using ^35^S-GAD_65_(143-585) compared with ^35^S-GAD_65_(1-585) and ^35^S-GAD_65_(96-585). Specificity for diabetes was improved compared with ^35^S-GAD_65_(1-585), similar to ^35^S-GAD_65_(96-585). Therefore, we focused on GAD_65_(96-585) for comparison with LIPS assays.

The large population, comprised of recent-onset patients and high- and low-risk FDRs from the well-characterised BOX study, with up to 30 years of follow-up, strengthened this study. Although samples were pre-screened by RBA, a large cohort of GADA negative FDRs was included to overcome this selection bias.

Overall, fewer low risk-relatives were positive for GADA(1-585) when measured by LIPS than RBA although future studies in an independent cohort are merited. Nluc-GAD_65_ antigens were fused to the N-terminus of the GAD_65_ constructs which may explain why assay performance was improved in the Nluc-GAD_65_(1-585) compared with the ^35^S-GAD_65_(1-585) test. Primary diabetes-associated epitopes of GAD_65_ are located in the middle and C-terminal domains(15-20) while minor N-terminal reactivity results from epitope spreading (21, 22). Nluc enzyme fusion may have obscured non-specific, linear epitopes and supported the stability and/or solubility of the antigens accounting for the enhanced performance observed.

RBAs have dominated islet autoantibody measurement for two decades but are disadvantaged by the high cost, short shelf life, tight regulation and environmental impact. Although alternative, sensitive and specific, non-radioactive methods are available for GADA measurement, these too have limitations, including large serum requirements and/or the need for specialist equipment and reagents(7, 23). The LIPS format was designed to be a simple replacement for the harmonised fluid-phase GADA RBA, using common techniques based on the precipitation of autoantibodies bound to cognate tracer antigens. Laboratories already set up to perform RBAs can easily adopt this method using widely available equipment and reagents. Luciferase-tagged antigens are safe and can be produced in-house with potentially long half-lives, giving greater control over label variability and eliminating reliance on radiolabels. The protocol can be completed within one working day and has the lowest serum requirement of all the widely available tests (2µl for testing in duplicate). This is critical for high-throughput testing of low-volume samples, for instance capillary blood collection for general population screening(24, 25). We also demonstrated the flexibility of LIPS, which can use a range of GAD antigens to facilitate with epitope analysis.

These assays were among the top performers in the 2017 IASP workshop. Future intervention trials and natural history studies may benefit from using this method and/or truncated antigens to measure GADA for identifying high-risk subjects.

## Supporting information

ESM 1a ESM 1b

## Acknowledgements

The BOX Study Group is comprised of Drs Chitrabhanu Ballav, Atanu Dutta, and Michelle Russell-Taylor, Bucks Healthcare Trust, UK; Dr Rachel Besser, Oxford University Hospitals Trust UK, UK; Drs James Bursell and Shanthi Chandran, Milton Keynes University Hospital, UK, Dr Sejal Patel, Wexham Park Hospital, UK; Drs Anne Smith and Manohara Kenchaiah, Northampton General Hospital, UK; Dr Gomathi Margabanthu, Kettering General Hospital, UK; Drs Foteini Kavvoura and Chandan Yaliwal, Royal Berkshire Hospital, UK.

## Author Contributions

RCW, AJKW (deceased) and KMG contributed to the original idea and designed the study. RCW, SLG, BTG, AEL and MA-MC designed and performed data collection. DL, CB and VL created the luciferase tagged GAD constructs. RCW, SLG, AEL, KMG and AJKW performed data analysis and wrote the manuscript. All authors contributed to data interpretation, manuscript revision and approved the final manuscript. AEL and KMG are responsible for the integrity of the work as a whole. RCW and SLG are joint first authors.

## Prior Presentation

Wyatt R.C., Liberati D., Brigatti C., Grace S.L., Gillard B.T., Long A.E., Shoemark D., Marzinotto I., Gillespie K.M., Achenbach P., Piemonti L., Lampasona V., Williams A.J.K. (2018) “A luciferase-based immunoprecipitation system (LIPS) assay using a truncated glutamate decarboxylase (GAD) tracer improves the specificity of GAD autoantibody measurement” (Poster presentation) *The Diabetes UK Professional Conference*, March 14-16, London, UK. RC Wyatt, D Liberati, C Brigatti, B Gillard, DK Shoemark, KM Gillespie, P Achenbach, L Piemonti, V Lampasona, AJK Williams (2017) “Measurement of autoantibodies to glutamate decarboxylase (GAD) using a truncated luciferase tracer offers improved specificity for type 1 diabetes.” (Poster presentation) *Immunology of Diabetes Society 15^th^ International Congress*, January 19-23, San Francisco, USA.

## Data availability statement

Data supporting conclusions reported in this manuscript are available upon reasonable request.

## Ethics Statement

The Bart’s Oxford study is currently approved by the South Central-Oxford C. National Research Ethics Committee. Participants provided informed, written consent and the study was performed according to the principles of the Declaration of Helsinki.

