## Supplementary material for "Improved specificity of glutamate decarboxylase 65 autoantibody measurement using luciferase-based immunoprecipitation system (LIPS) assays": ESM 1a ESM 1b

### ESM 1

(a)

| | Patients (n=154) | | GADA Positive: Progressors (n=69) | | GADA Positive: Multiple islet autoantibody positive (n=36) | | GADA Negative: Progressors (n=32) | |
|---|---|---|---|---|---|---|---|---|
| | **Median DK units/ml (range)** | **Positives (n)** | **Median DK units/ml (range)** | **Positives (n)** | **Median DK units/ml (range)** | **Positives (n)** | **Median DK units/ml (range)** | **Positives (n)** |
| $^{35}$S-GADA (1-585) | 121.0 (0-1340.5) | 125 (81%) | 237.8 (0-1034.2) | 65 (94%) | 330.8 (2.2-843.8) | 33 (92%) | 0.3 (0-10.6) | 0 |
| $^{35}$S-GADA (96-585) | 106.8 (0-1447.4) | 125 (81%) | 253.6 (0-1050.8) | 63 (91%) | 323.7 (0-1024.0) | 31 (86%) | 1.0 (0-12.9) | 1 (3%) |
| Nluc-GADA (1-585) | 103.4 (0-1859.0) | 129 (84%) | 327.8 (0-1732.1) | 64 (93%) | 265.4 (0.0-1138.8) | 34 (94%) | 0 (0-12.9) | 1 (3%) |
| Nluc-GADA (96-585) | 78.62 (0-1159.9) | 116 (75%) | 341.85 (0-1012.2) | 57 (89%) | 265.4 (0-927.0) | 31 (86%) | 0 (0-22.17) | 1 (3%) |

(b)

| | GADA +ve Non-progressors (n=173) | | GADA –ve Non-progressors (n=430) | |
|---|---|---|---|---|
| | **Median DK units/ml (range)** | **Positives (n)** | **Median DK units/ml (range)** | **Positives (n)** |
| $^{35}$S-GADA (1-585) | 49.8 (0-1141.8) | 156 (90%) | 0 (0-40.8) | 12 (3%) |
| $^{35}$S-GADA (96-585) | 29.8 (0-1054.2) | 108 (62%) | 0.6 (0-32.9) | 6 (1%) |
| Nluc-GADA (1-585) | 20.1 (0-1919.8) | 112 (65%) | 0 (0-208.0) | 17 (3%) |
| Nluc-GADA (96-585) | 4.5 (0-1130.0) | 74 (43%) | 0 (0-239.10) | 10 (2%) |
